# Reducing contacts to stop SARS-CoV-2 transmission during the second pandemic wave in Brussels, Belgium

**DOI:** 10.1101/2020.12.23.20248795

**Authors:** Brecht Ingelbeen, Laurène Peckeu, Marie Laga, Ilona Hendrix, Inge Neven, Marianne A. B. van der Sande, Esther van Kleef

**Affiliations:** Department of Public Health, Institute of Tropical Medicine, Antwerp, Belgium; Department of Infectious Disease Prevention and Control, Common Community Commission, Brussels-Capital Region, Brussels, Belgium; Julius Center for Health Sciences and Primary Care, Utrecht University, the Netherlands

## Abstract

**Background:** Reducing contacts is a cornerstone of containing SARS-CoV-2. We evaluated the effect of physical distancing measures and of school reopening on contacts and consequently on SARS-CoV-2 transmission in Brussels, a hotspot during the second European wave.

**Methods:** Using SARS-CoV-2 case reports and contact tracing data during August-November 2020, we estimated changes in the age-specific number of reported contacts. We associated these trends with changes in the instantaneous reproduction number *R*_*t*_ and in age-specific transmission-events during distinct intervention periods in the Brussels region. Furthermore, we analysed trends in age-specific case numbers, pre- and post-school opening.

**Findings:** When schools reopened and physical distancing measures relaxed, the weekly mean number of reported contacts surged from 2.01 (95%CI 1.73-2.29) to 3.04 (95%CI 2.93-3.15), increasing across all ages. The fraction of cases aged 10-19 years started increasing before school reopening, with no further increase following school reopening (risk ratio 1.23, 95%CI 0.79-1.94). During the subsequent month, 8.9% (67/755) of infections identified were from teenagers to other ages, while 17.0% (131/755) from other ages to teenagers. Rt peaked mid-September at 1.48 (95%CI 1.35-1.63). Reintroduction of physical distancing measures reduced reported contacts to 1.85 (95%CI 1.78-1.91), resulting in Rt dropping below 1 within 3 weeks.

**Interpretation:** The second pandemic wave in Brussels was the result of increased contacts across all ages following school reopening. Stringent physical distancing measures, including closure of bars and limiting close contacts while schools remain open, reduced social mixing, in turn controlling SARS-CoV-2 transmission.

**Funding:** European Commission H2020. GGC Brussel.

## Introduction

Belgium has been particularly hard hit by the COVID-19 pandemic. The country reported the highest number of deaths per capita and near highest number of cases per capita worldwide. During Europe’s ‘second pandemic wave’ the country was again the worst-affected country in Europe in per capita case numbers and deaths^1^. Belgium loosened physical distancing measures at a moment when case numbers were rising^2^. Brussels, Belgium’s capital, was ahead of the rest of Belgium to observe a steep increase in cases and to step up preventive measures^3^.

These interventions, involving physical distancing measures, target the reduction of person-to-person contact in order to reduce the number of occasions a virus can be transmitted. While close-contact interactions are considered to play a key role in SARS-CoV-2 transmission, infection rates are generally assumed to proportionally in- or decrease with changes in (age-specific) number of contacts^4^. Hence, previous modelling studies estimated the effect of physical distancing by evaluating the impact of changes in the general population’s contact patterns on *R*_*0*_ during and post lockdown^5,6^.

In this study, using operational data, we describe the effect of physical distancing measures and the closure of schools on contact patterns and SARS-CoV-2 transmission. We analysed trends in reported contacts during distinct intervention periods in the Brussels region, and associated these trends with estimated transmission patterns, and age-specific case numbers over time during Brussels’ second pandemic wave.

## Methods

We used data generated by the test and contact tracing system of the Brussels region between 1 August and 12 November 2020, to deduct contact and transmission patterns, and official data on COVID-19 case reports for Brussels made available via the Belgian institute for health, Sciensano, to assess age-specific trends in case numbers^3^.

### Data processing

In May 2020, Belgium implemented a phone- and field agent-based contact tracing system. SARS-CoV-2 PCR-positive cases and their contacts were identified and requested to self-isolate. High-risk (close) contacts, defined as physical or cumulative 15 minutes non-physical contact within 1.5m from 2 days before to 7 days after onset of symptoms of a confirmed SARS-CoV-2 case, were recommended to undergo SARS-CoV-2 PCR testing, regardless of symptoms. For contacts aged 0-6 years, and from October 21 onwards across all ages, testing was restricted to symptomatic individuals only^7^. In primary schools, pupils and teachers in the same class of a confirmed case were considered low-risk contacts, therefore did not require testing, except if presenting symptoms. In secondary schools, the regular high-risk contact definition and testing criteria are applied. Pseudonymised data on SARS-CoV-2 contacts generated by the contact tracing system were linked to SARS-CoV-2 case data (including age) using a unique identifier based on first and last name. Homonyms that resulted in duplicates with the same unique identifier were excluded from the dataset. Hence, we identified contacts that tested SARS-CoV-2 positive within 3 weeks after the reported date of contact with an index case, generating a database with transmission events between primary (index) and secondary cases (contacts).

We identified when changes in physical distancing measures were introduced by the national and regional governments, distinguishing six distinct time periods with different combinations of physical distancing measures and school closure, which we refer to as *intervention periods* (Table 1).

**Table 1.** Physical distancing measures and SARS-CoV-2 testing policy changes during July-November 2020 in the Brussels region

| Intervention | Start | Period |
| --- | --- | --- |
| Cafés and restaurants may remain open until 1 a.m. and can take maximum 10 people per group | 8 June |  |
| Sports allowed in groups of maximum 50 people | 8 June |  |
| Maximum 5 close contacts* per week | 30 July | 1 |
| Reopening primary and secondary schools | 1 Sep | 2 |
| Restart universities at 50% to 75% room occupancy, with masks | 14 and 21 Sep |  |
| Limit on number of close contacts suspended | 30 Sep | 3 |
| Quarantine for high risk contacts reduced from minimal 10 days to 7 days (if two negative tests) | 30 Sep |  |
| Maximum 3 close contacts per week | 6 Oct | 4 |
| Recommended teleworking | 6 Oct |  |
| Bars and cafés closed at 23h | 6 Oct |  |
| Bars and cafés closed | 8 Oct |  |
| Universities restrict seat occupancy to 20% | 19 Oct |  |
| Testing restricted to symptomatic suspected SARS-CoV-2 cases (except for healthcare workers) | 21 Oct |  |
| Quarantine for high risk contacts extended to 10 days | 21 Oct |  |
| Restaurants closed | 26 Oct | 5 |
| Maximum 1 close contact per person and private gathering with max. 4 people (excluding <12 year olds) | 26 Oct |  |
| Curfew between 10 pm and 6 am | 26 Oct |  |
| Teleworking "becomes the rule" | 26 Oct |  |
| Indoor sports prohibited (except <12 year olds) | 26 Oct |  |
| Universities switch to on-line learning (VUB) | 26 Oct |  |
| Maximum 1 close contact per household | 2 Nov | 6 |
| Mandatory teleworking | 2 Nov |  |
| Non-essential shops closed; professions involving physical contact or gatherings suspended | 2 Nov |  |
| Extended autumn school holiday (31 Oct-15 Nov) | 31 Oct |  |

\*Closer contact for more than 15 minutes with a person who is not part of your household, without keeping a distance of 1m50 and not wearing a mask, excluding children under 12 years old; Sources: https://www.commissioner.brussels/updates-covid-19; https://covid-19.sciensano.be/nl

### Data analyses

#### Changes in contact patterns over time

We computed the mean number of contacts reported per case per week, overall and by age group. We compared differences in sample mean and confidence intervals for weekly contacts at the start and end of each intervention period assuming normality after visual inspection. To visualise and describe changes in daily contact patterns over time, we fitted a segmented linear regression allowing for a step and slope changes between distinct intervention periods (Supplementary material).

Transmissibility as well as contact patterns are known to vary by age. The age of contacts was only limitedly listed. Hence, we were unable to construct social contact matrices, capturing contact patterns between age-groups. Social contract matrices with a per contact infectivity value (often denoted as q) allow for calculation of the next generation matrix (NGM) describing the number of potential transmission events per individual per age group. The dominant eigen value of this NGM gives an estimate of the basic reproduction number *R*_*0*_ as a metric for transmission^4^, and has previously been used as a metric to assess the impact of changes in physical contacts^5,6^.

As alternative, we characterised changes in SARS-CoV-2 transmission over time, by estimating the non-age specific instantaneous reproduction number *R*_*t*_, i.e. the mean number of secondary cases that would arise from a primary case on a given day, during our study period. Employing established methods^8^, we derived *R*_*t*_ from the daily number of reported cases, assuming an uncertain serial interval distribution (i.e. drawn from multiple truncated normal distributions with mean 5.19 days, 95%CI 4.37-6.02)^8,9^. We set a seven-day sliding window; bootstrapping was used to obtain robust confidence intervals. The impact of changes in contact patterns was estimated by quantifying the relative change in *R*_*t*_.

#### Impact school opening

To further investigate the impact of changes in contact patterns on SARS-CoV-2 transmission dynamics, we quantified the relative frequency of transmission events between age groups over time. We extended the period to 30 November, to allow for the evaluation of transmission-events after the extended autumn holidays. We characterised the (change in) degree of intra- and intergenerational transmission events during periods where contact patterns changed, including post-opening of schools.

Finally, we describe changes in the proportion of daily reported cases among secondary school-aged children (10-19 years old) in the months pre- and post-school opening (August to September), comparing segmented and non-segmented Poisson regression models (Supplementary material). We hypothesised that, if secondary schools acted as a predominant driver of SARS-CoV-2 transmission during Brussels’ second wave, the fraction of 10-19 year olds among all reported cases would change first following school opening, before extending to other generations. We adjusted the periods for reporting delays by including a lag between exposure and case report (4 days) and compared model fits based on AIC assuming different time trends following school opening.

All analyses were done in R version 4.0.2 (R Foundation for Statistical Computing, Vienna, Austria; packages ‘EpiEstim’, ‘stats’, ‘ggplot2’). Scripts are accessible on a GitHub repository: https://github.com/ingelbeen/covid19bxl. The study was approved by the Institutional Review Board of the Institute of Tropical Medicine and the Ethics committee of the Antwerp University Hospital.

## Results

From 1 August to 12 November 2020, the Brussels region reported 63,838 SARS-CoV-2 confirmed cases (5.2% of its population) from 415,412 SARS-CoV-2 PCR tests performed. The daily number of confirmed cases peaked on October 20 with 2,950 cases reported (figure 1). SARS-CoV-2 test positivity was highest among 20-29 years olds (7.4%, 13,436/181,940), and decreased with age, with 4.3% positive (4,913/114,637) among 70+ year olds (Supp Fig 1). A total of 52,484 cases were referred for contact tracing. Among these cases, 24,166 (46.0%) reported at least one contact, 61,754 in total. Matching operational case and contact databases resulted in a final 19,194 cases with recorded age and 51,177 contacts. The time between the last reported contact and contact tracing was median 2 days (interquartile range 0-5 days). Until 30 November, we traced back 2,443 reported contacts that tested SARS-CoV-2 positive within 3 weeks, yielding primary-secondary case pairs, 2,387 with age recorded.

**Fig 1.**
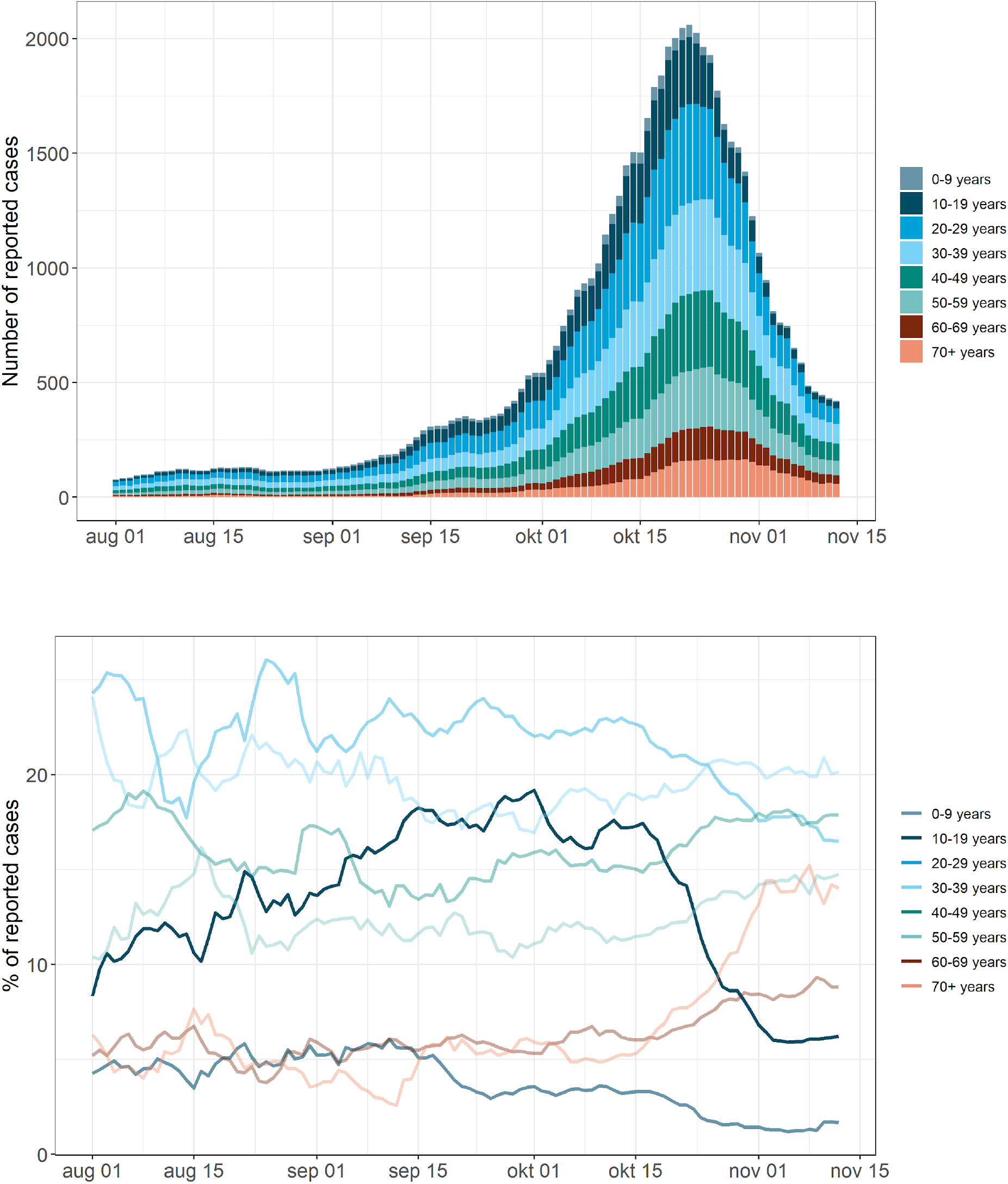
7-day moving average of SARS-CoV-2 confirmed cases reported in Brussels region between August 1 and November 12, 2020. A. Number of cases; B. Percentage of reported cases per age group over time.

### The effect of physical distancing measures on the number of reported contacts

August saw a wide variation in the number of reported contacts per case and the percentage of cases reached compared to the following months (Supp Fig 2). September noted a significant increase in the mean number of reported contacts, from 2.01 (95%CI 1.73-2.29) in the last week of August pre-school opening (period 1), to 2.83 (95%CI 2.59-3.06) in the first week of September (period 2, Fig 2, Supp Fig 2). We found no change in the number of reported contacts when the restriction on the number of close contacts was suspended on September 30 (period 3), plateauing at mean 3.04 (95%CI 2.93-3.15). In the fourth intervention period, involving the restriction to 3 close contacts and the closure of bars on October 6 and 8, resulted in a gradual 21% decrease in reported contacts from mean 2.81 (95%CI 2.74-2.89) in the first week to 2.21 (95%CI 2.16-2.25) before contacts were further limited on 26 October. A week into the 5^th^ period with a limit of one close contact and a closure of restaurants and sports facilities, a further decrease was observed to 1.94 reported contacts (95%CI 1.90-1.99), i.e. 45% decrease compared to September 30. When also shops were closed, telework became mandatory, and schools started the autumn break, the mean number of reported contacts stabilised at 1.85 (95%CI 1.78-1.91).

**Fig 2.**
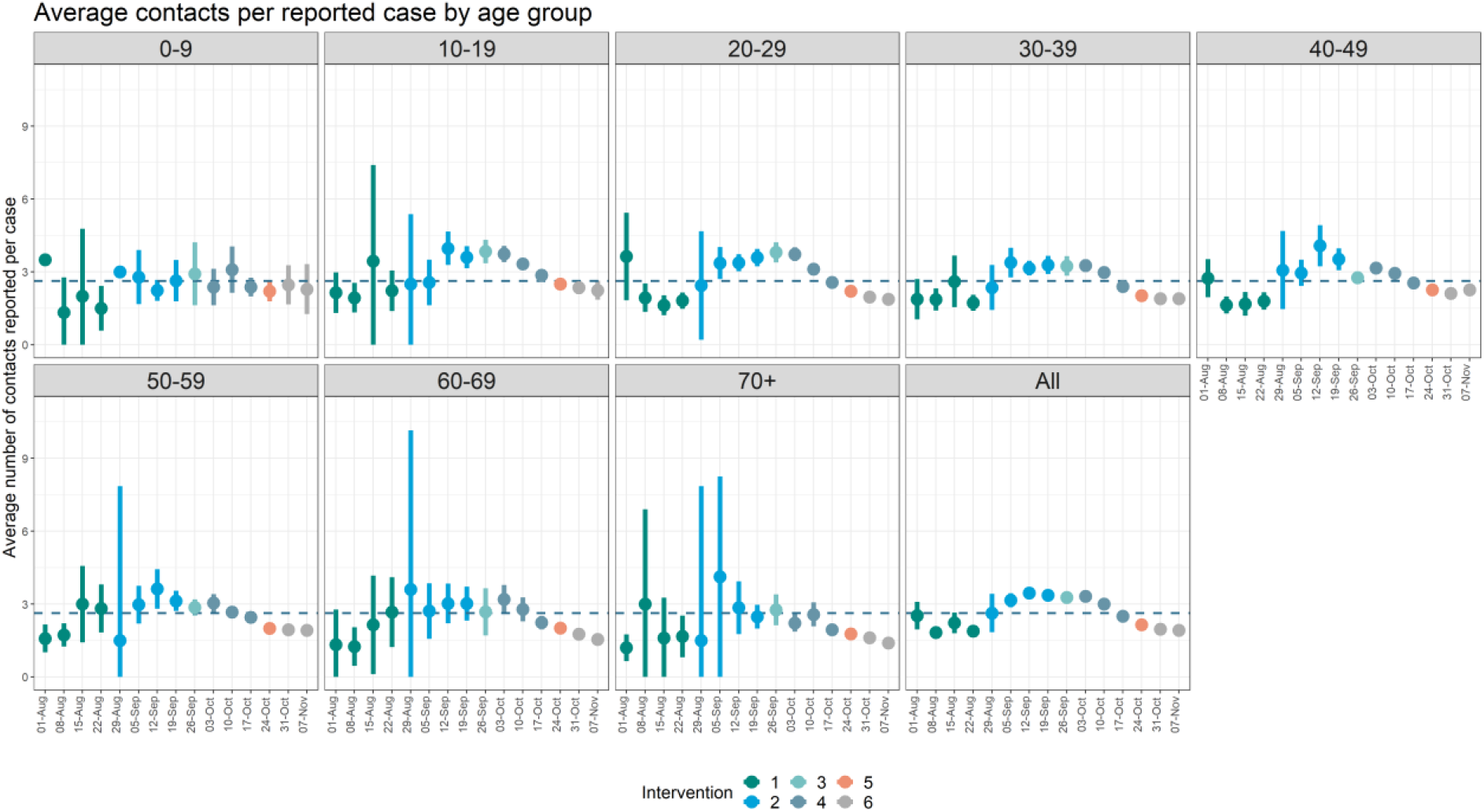
Weekly mean number of contacts reported per SARS-CoV-2 case (excluding cases not reporting any contacts) and 95% confidence intervals by age group. Dotted line represent the mean number of cases for all age groups the week of school reopening. Of note, weeks follow 7 day intervals from 1st of August. Hence, start of the weeks do not correspond with the starting dates of intervention periods. Colours merely indicate the week during which the respective interventions started and ended. For trends in daily estimates and exact timings, see supp Fig 3. **Green =** schools open & 5 close contacts allowed; **Blue =** schools open & limit close contacts suspended; **Light blue** = schools open, bars closed & 3 close contacts allowed; **Grey** = schools open, bars & restaurants closed, curfew, indoor sports prohibited & 1 close contacts allowed; **Orange** = schools closed, mandatory teleworking, non-essential shops closed and all of the above. For readability, the wide confidence intervals of the observation of the first week for age 0-9 were removed.

10-19 year olds reported overall the highest number of contacts during our study period (3.11, 95%CI 3.01-3.21); adults aged 70+ years reported the lowest number (2.05, 95%CI 1.93-2.18). However, over time, changes in the number of contacts following changes in physical distancing measures were similar across age-groups (Fig 2).

### Effect of the number of reported contacts on SARS-CoV-2 spread

*R*_*t*_ ranged from 1.66 (95%CI 1.36-1.93) on August 2 to 0.56 (95%CI 0.50-0.62) on 11 November. *R*_*t*_ peaked September 17, at 1.48 (95%CI 1.35-1.63) following the September-October surge in case numbers (Fig 3). After the limitation to 3 contacts and closure of bars (periods 4), *R*_*t*_ decreased with 42% to 0.82 (95%CI 0.79-0.85) before the extended autumn holidays, i.e. the first week into the period of a further limit to one close contact, closure of restaurants and sport facilities (period 5), dropping below 1 on 29 October.

**Fig 3:**
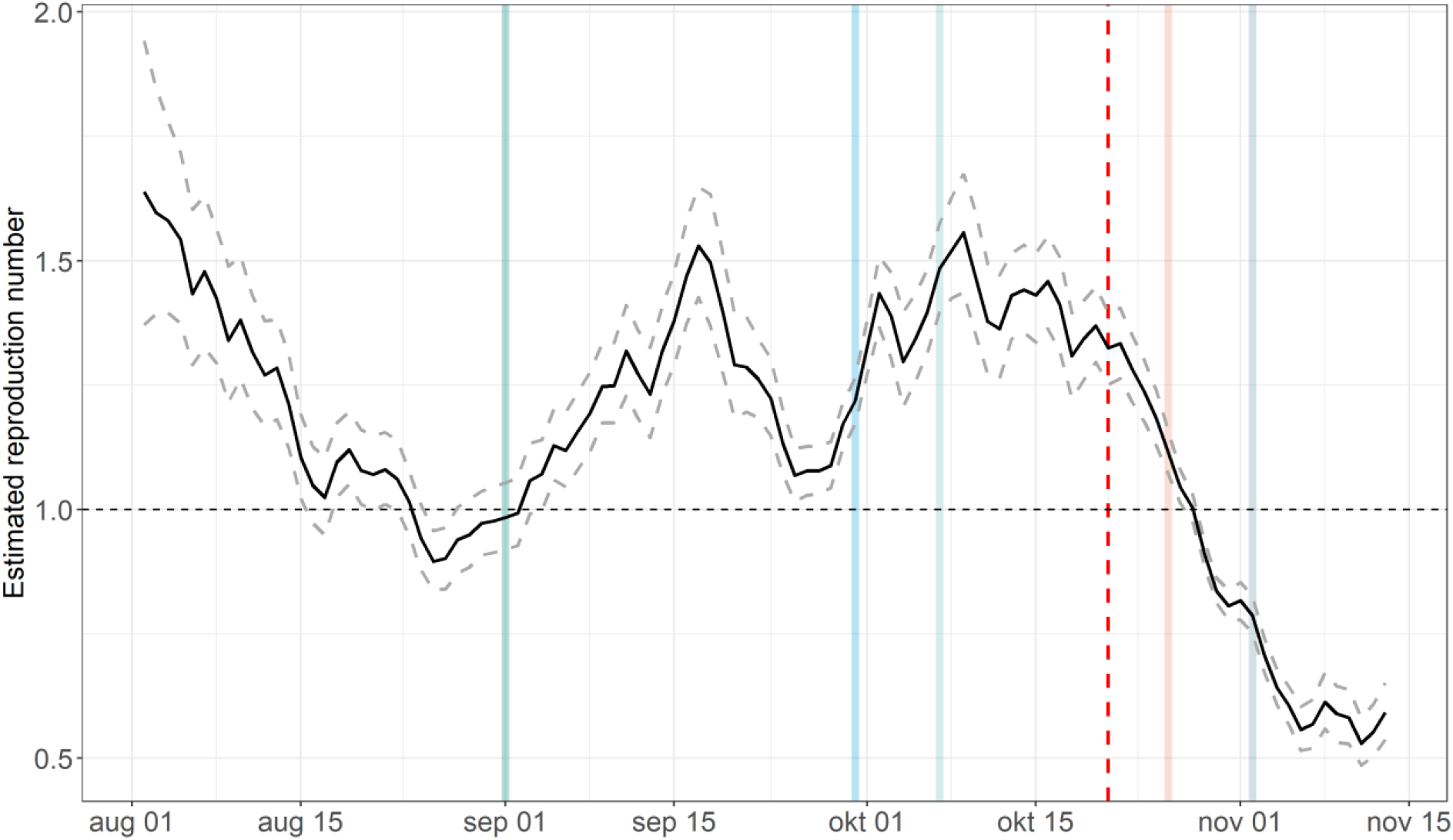
Estimated instantaneous reproduction number (*R*_*t*_) based on daily reported cases and a mean 5.2 day serial interval (95%CI 4.4-6.0; Rai et al) using the EpiEstim R package. After October 21 (the dashed red line) asymptomatic contacts were excluded from SARS-CoV-2 testing. Vertical lines represent the intervention periods. **Green =** schools open & 5 close contacts allowed; **Blue =** schools open & limit close contacts suspended; **Light blue** = schools open, bars closed & 3 close contacts allowed; **Orange** = schools open, bars & restaurants closed, curfew, indoor sports prohibited & 1 close contacts allowed; **Grey** = schools closed, mandatory teleworking, non-essential shops closed and all of the above.

### Age-specific transmission patterns

Among 2,387 identified primary-secondary case pairs, transmission within the same age group (33.0%, 797/2,387) was predominant across age groups throughout all time periods. From November 4 onwards, after introducing stringent physical distancing measures and during the extended autumn holidays, intrageneration transmission was highest at 39% (63/160). Infections from 10-19 year olds were seldom recorded in August and November when schools were closed, but testing of this group was low at these times as well (Fig 4, Supp Fig 3). After schools reopened, transmission between all age-groups became more apparent, reducing again after November 4, which saw relatively more transmission-events within older age-groups (50+). Furthermore, in the month after reopening schools, 8.9% (67/755) of infections were from 10-19 year olds to other age groups and 17.0% (131/755) from other age groups to 10-19 year olds.

**Fig 4.**
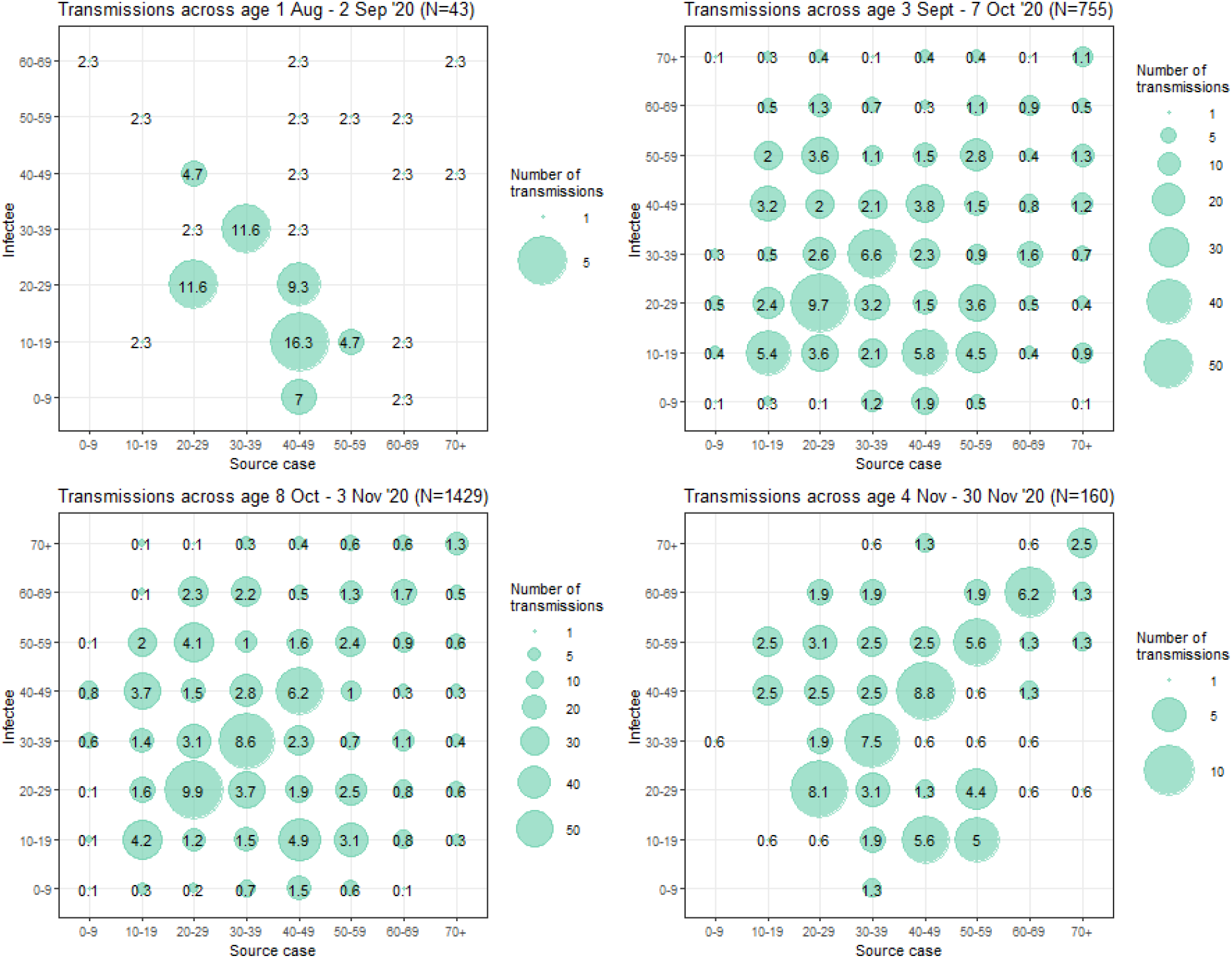
Transmission matrix between index and secondary cases of all identified transmission events. A. Pre-school opening (1 August to 2 September 2020); B. First month post-school opening (3 September to 7 October 2020); C. Second month post-school opening until schools closed for an extended autumn school holiday (from October to 3 Nov 2020). D. Period of extended autumn school holiday and two-weeks after (4 to 30 November 2020).

### Age-specific trends in SARS-CoV-2 reported cases

Up to October 20, a notable increase in reported SARS-CoV-2 was observed among 10-19 year olds (Figure 1B), coinciding with a testing rate increase in this age class over time (spearman rank correlation coefficient = 0.74, p-value<0.000, Supp Fig 2). At the time schools reopened (1 September), we observed no significant change in the fraction of 10-19 year olds among all diagnosed cases (risk ratio 1.23, 95%CI 0.79-1.94, Supp Fig 5). When asymptomatic contacts were excluded from SARS-CoV-2 testing (October 21 onwards), the fraction of 10-19 year olds fell from 16.9% of the cases (3478/20,535 during 2 preceding weeks) to 9.9% (2214/22,330 during 2 following weeks, Fig 1B). The fraction of adults aged 70 or more, who tested positive, increased by mid-October, from 5.2% (727/13872) during the two first weeks of October to 13.8% (1574/11430) in the first two of November (Fig 1B).

## Discussion

September 2020 saw a persistent increase in SARS-CoV-2 cases in the Brussels region following increased social mixing across all ages. Stringent physical distancing measures were introduced one month after an alarming persistent increase in *R*_*t*_ (6 October). These initial measures (a limit to three close contacts, a curfew, a closure of bars, and recommended teleworking) reduced reported contacts to less than two after three weeks (a 45% decrease), resulting in an *R*_*t*_ <1 on 29 October. This suggests stringent physical distancing measures can sufficiently reduce social mixing to control transmission, even without closing schools or full lockdown.

In contrast to the first pandemic wave, schools remained open throughout the second wave. There is general consensus that children attending primary school contribute little to transmission^10,11^. In contrast, the role of teenagers and secondary schools is still much debated^12^. Teenagers can transmit and show comparable viral load to adults^13^. Nonetheless, several studies indicated either lower susceptibility or a higher proportion of asymptomatics among children which might result in fewer secondary infections from younger individuals^11,14–16^. Modelling studies investigating the role of secondary schools have shown school closures can help alter transmission-dynamics – albeit insufficiently for control and based on data from the first months of the pandemic with limited preventive measures in schools^16^. Others described transmission in schools, although without considering its potential contribution to SARS-CoV-2 dynamics in the general population^17,18^. Our findings confirm transmission among and from teenagers, with intergenerational transmission apparent following school opening. Nonetheless, their relative role was limited: transmission events from 10-19 year olds to other age groups remained inferior compared to those from adults when schools were open, and the fraction of cases among 10-19 year olds did not significantly change after school reopening. This was at a time testing among teenagers was extensive, reducing the likelihood of their infections remaining undetected. An increase preceding school reopening was largely explained by an increase in testing rates among this population (Supp Fig 3).

After school reopening, the number of reported contacts increased across all age groups. Transmission among same-age groups predominated, in line with extensive contact tracing and testing data from India^19^, which was related to both a higher frequency of contact and a higher per-contact probability of infection with individuals of a similar age^19,20^.

With testing and contact tracing overwhelmed by increasing case numbers, testing and tracing become nearly ineffective in outbreak control^21^. Physical distancing measures are then an effective yet costly intervention to slow SARS-CoV-2 transmission^22–24^. In Brussels, even though epidemic growth could be slowed within three weeks through physical distance measures, reducing case numbers to such extent to allow effective testing and tracing proved to take longer. Planned, short periods of strict measures, so-called precautionary breaks, have been proposed as a more effective and less costly tool to control SARS-CoV-2 spread^25^.

We found epidemic growth to be delayed among older adults, similar to observations in other European countries, including Spain and the UK^26,27^. From October 15 onwards, the fraction of older adults among SARS-CoV-2 cases in Brussels started to increase (Fig 1B). The change in testing strategy excluding asymptomatic contacts from testing from October 21 can only partially explain this shift. The proportions of asymptomatic infections differ, but not to such extent between older and younger age groups^15^. SARS-CoV-2 transmission has shown to vary between age-groups and settings, revealing so-called superspreading events^28–30^. Individuals with social networks less linked to the general population, such as older adults – nursing home residents in particular – can disproportionately increase when a certain threshold of infections is reached in the general population^31^. This so-called percolation phenomenon may explain why older adults were less affected before the epidemic peak, but increased in case numbers thereafter. This emphasizes the importance of testing, and soon vaccinating, key persons that are likely to link older frail adults to the overall population, e.g. nursing home staff or healthcare workers.

To our knowledge, our study is the first to evaluate the role of physical distance measures on social mixing and SARS-CoV-2 control during the second pandemic wave in Europe, using operational data. Because testing was extensive across all ages except 0-6 year olds, including high-risk contacts with asymptomatic infections, age-specific transmission patterns are more robust than during the first months of the pandemic. Some precaution is warranted though when interpreting our findings. First, the number of high-risk contacts reported by cases in our study was lower (mean 2.0, 95% CI 1.8-2.0, in August) than what participants in a social mixing survey in Belgium reported (mean 3.5 during 27 July-10 August)^5^. This can partially be explained by limited recording or re-calling of low-risk contacts or context-specific accidental social contacts (e.g. public transport, bars), or could relate to individuals being reluctant in reporting all contacts. Yet, age-specific differences were comparable, suggesting our conclusions relying on reported trends over time, remain valid.

Linkage of contacts and cases resulted in a low fraction of cases that where a known contact, indicating high volumes of undetected transmission. National identification number based matching could have improved our linkage, but was only well recorded during periods of lower case volumes. Importantly, our cross-generation transmission events during November, coincided with a shift in testing to symptomatic cases as well as involvement of school doctors in contact tracing. With children and teenagers more frequently presenting without or with mild symptoms^32,33^, their lower involvement in transmission events during the period of an extended school holiday should be considered with care. This shift in testing could have resulted in an underestimation of *R*_*t*_ at the end of October. However, *R*_*t*_ continued to steadily decrease after the testing strategy change, suggesting a true drop in transmission-levels is likely.

In conclusion, using operational case and contact tracing data, we were able to evaluate the effect of physical distancing measures and school reopening on trends in age-specific contact patterns and SARS-CoV-2 transmission patterns. The intensity of the second pandemic wave in Brussels was a result of increased social mixing across all ages in absence of strict physical distancing measures. Reopening of schools coincided with an increase in SARS-CoV-2 transmission. Nonetheless, our data suggests this is likely the result of increased social mixing across all ages, rather than driven by SARS-CoV-2 transmission among children or teenagers, followed by spreading to other ages. Physical distancing measures, including a closure of bars and limiting close contacts, resulted in a rapid decrease in the reported number of contacts, which in turn led to reducing SARS-CoV-2 transmission.

## Data Availability

Data analysis scripts are accessible on a GitHub repository: https://github.com/ingelbeen/covid19bxl.
Case report data was available from the website of Sciensano, the Belgian health institute

## Acknowledgments

No specific funding was provided for this study, but authors were supported by grants from the European Union’s Horizon 2020 programme under Grant Agreement MOOD N° 874850 and from the Common Community Commission of Brussels-Capital Region. We thank the Common Community Commission for reaching out to collaborate and provide insight at several stages of the study, and David Hercot for helpful last-minute discussions. We thank the contact tracing team for data collection, and Sciensano for making age-specific case report and testing data open access.

## Supplementary material

### Description of regression models

To visualise and describe changes in contact patterns over time, we fitted a segmented linear regression allowing for step and slope changes between distinct intervention periods as follows: 

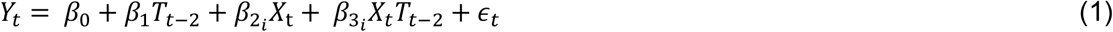

Where *Y*_*t*_ is the expected mean number of contacts on day *t. T*_t_ represents the day starting August 1, thus *β*_1_ can be interpreted as the underlying trend in contact patterns without any changes in interventions. *X*_*t*_ represents a dummy variable indexing the 6 distinct intervention periods *i*, with *β*_2_ and *β*_3_ representing the step and slope change in contacts following the introduction of interventions. We added a 2-day lag for delay between an at risk contact and reporting of that contact, based on the median number of days between the last reported contact and contact tracing.

We describe changes in the proportion of daily reported cases 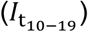 among teenagers in the months pre- and post-school opening (August to September), using Poisson regression with a log-link and offset term representing the total daily reported cases. 

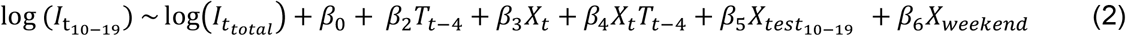

*T*_t_ represents the days from August until September, capturing the underlying trend pre-school opening, *X*_*t*_ represents a dummy variable indexing 0 and 1 before and after school opening respectively. We adjusted the periods for reporting delays by including a lag between exposure and case report (4 days). The daily number of tests performed among teenagers was accounted for and depicted by 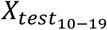 as well as whether the case was reported positive during the weekend *X*_*weekend*_. We compared model fits using Akaike Information Criterion (AIC), assuming different time trends following school opening (i.e. no, vs a step vs a step and slope change). Models with and without adjustment for school provided similar fits (AICs of 364.9, 363.1, and 362.4 for a model with a step and slope change, a step change only and no change at all respectively). Of note, models with and without testing showed similar fits, while 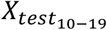 proved highly correlated with time.

## Supplementary figures

**Supp Fig 1A.**
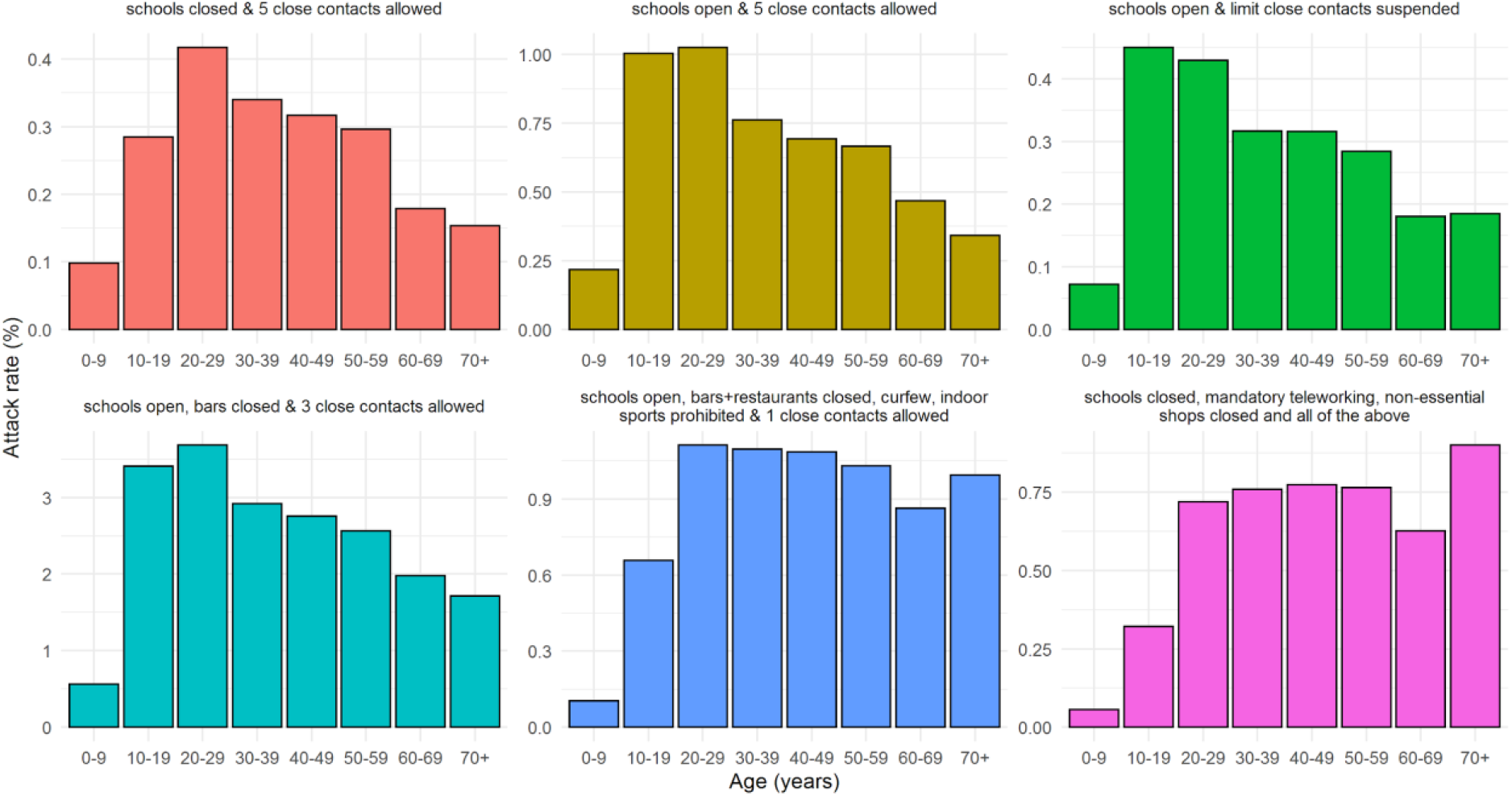
Percentage of the population which was SARS-CoV-2 confirmed by age group and by period of physical distancing measures. Source population numbers: https://statbel.fgov.be/en/themes/population/structure-population

**Supp Fig 1B.**
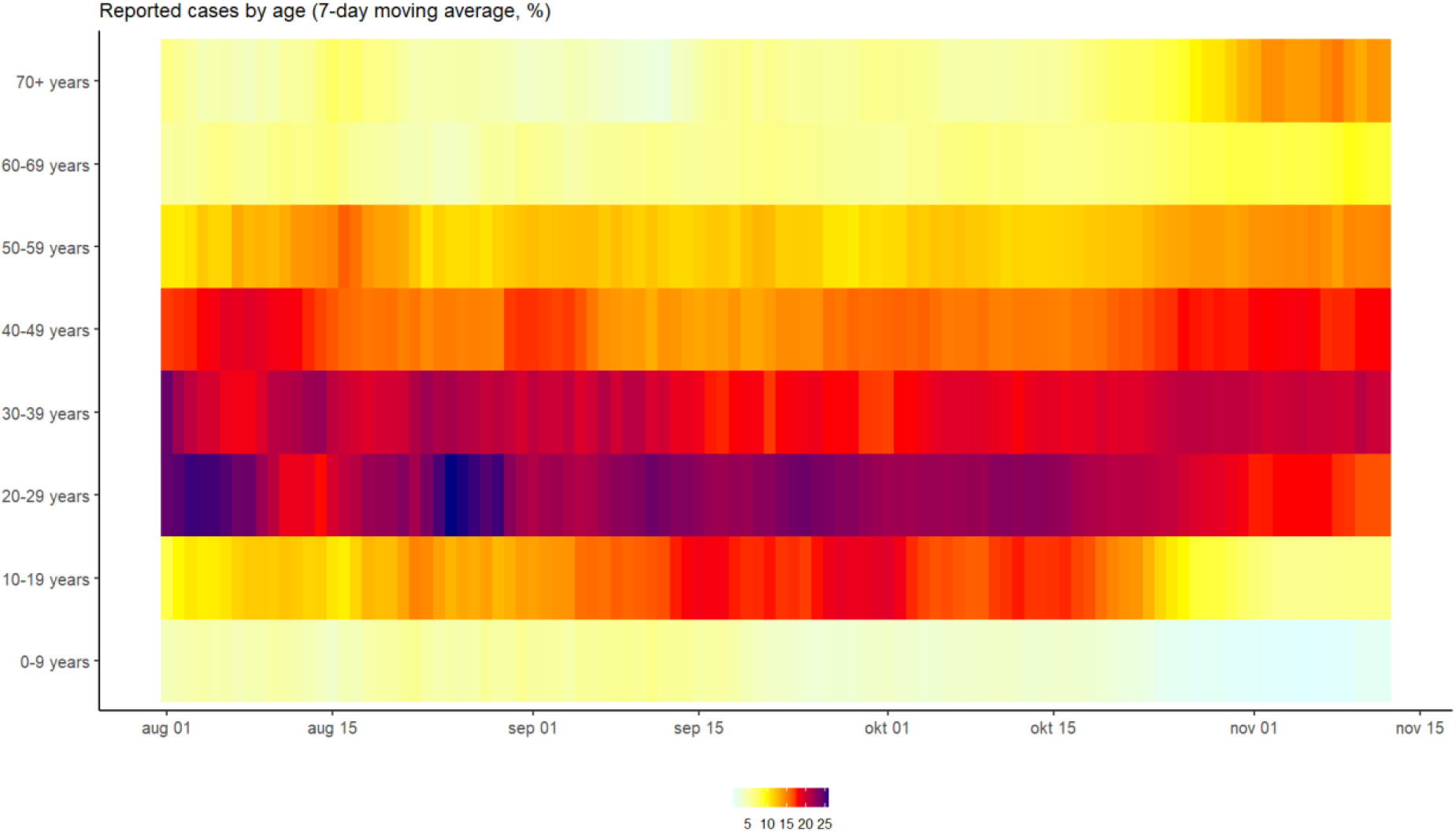
Percentage of the population which was SARS-CoV-2 confirmed by age group and by period of physical distancing measures. Source population numbers: https://statbel.fgov.be/en/themes/population/structure-population

**Supp Fig 2.**
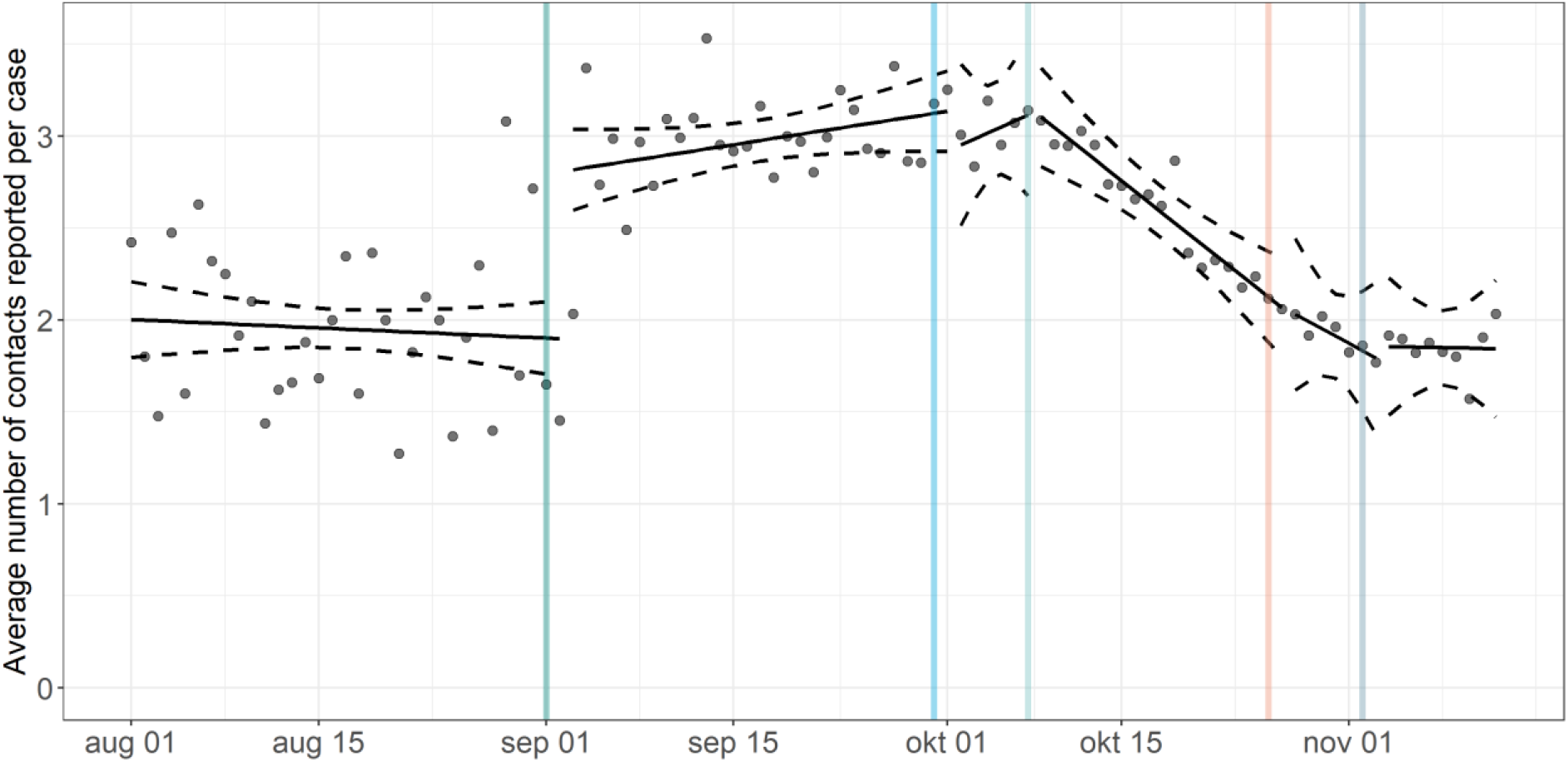
Daily mean number of contacts reported per SARS-CoV-2 case (excluding cases not reporting any contacts), with fitted estimated linear trends and 95% confidence intervals, using segmented linear regression with an interaction term for date and intervention periods, allowing for a step change. Lines are plotted as discontinuous for readability. The start of each segment in the linear regression is corrected for the median two days between the last reported contact and the interview. Vertical lines represent the intervention periods. **Green =** schools open & 5 close contacts allowed; **Blue =** schools open & limit close contacts suspended; **Light blue** = schools open, bars closed & 3 close contacts allowed; **Orange** = schools open, bars & restaurants closed, curfew, indoor sports prohibited & 1 close contacts allowed; **Grey** = schools closed, mandatory teleworking, non-essential shops closed and all of the above.

**Supp Fig 3.**
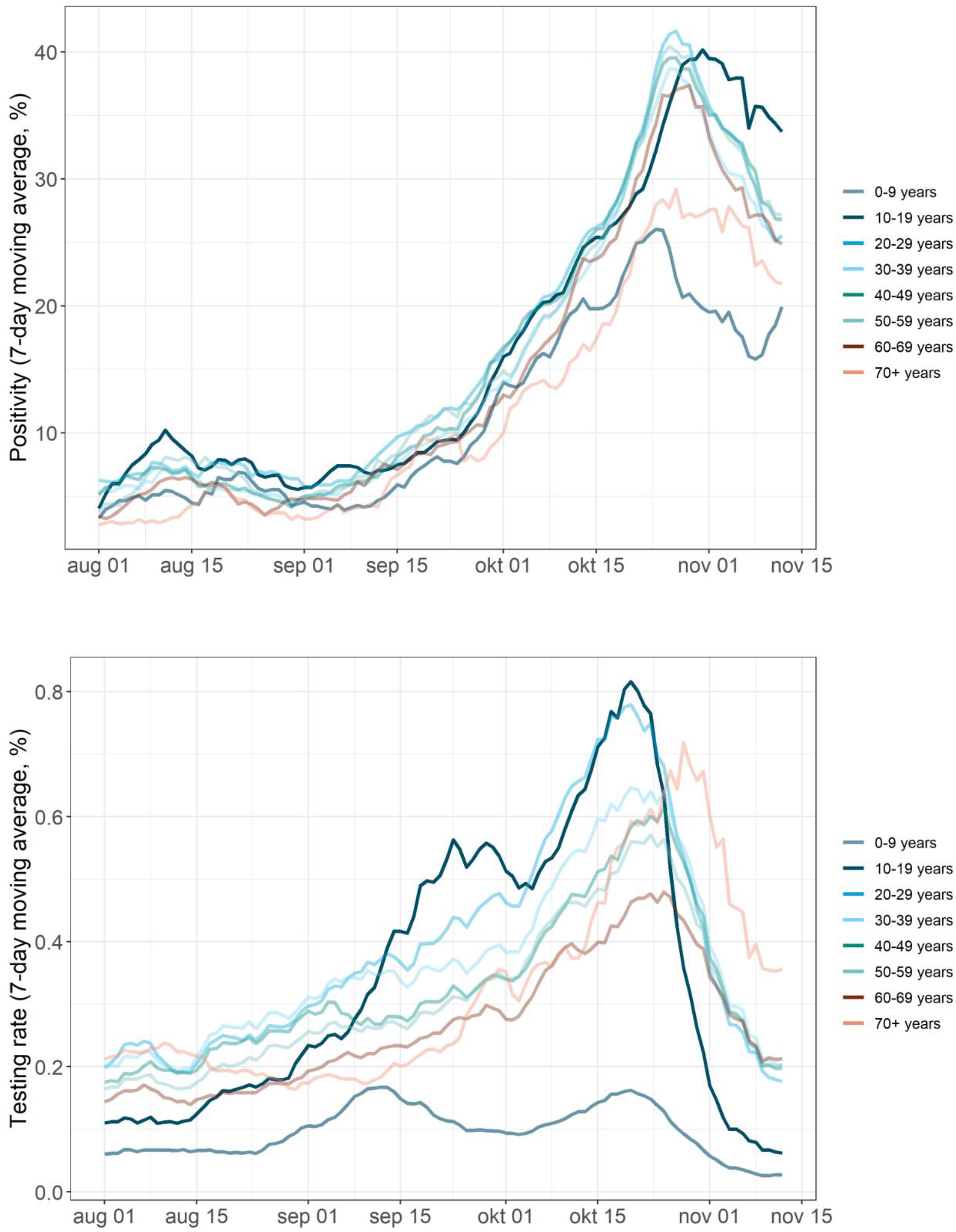
SARS-CoV-2 testing rate by age group in the Brussels region. Source: Sciensano and https://epistat.wiv-isp.be/covid/

**Supp Fig 4.**
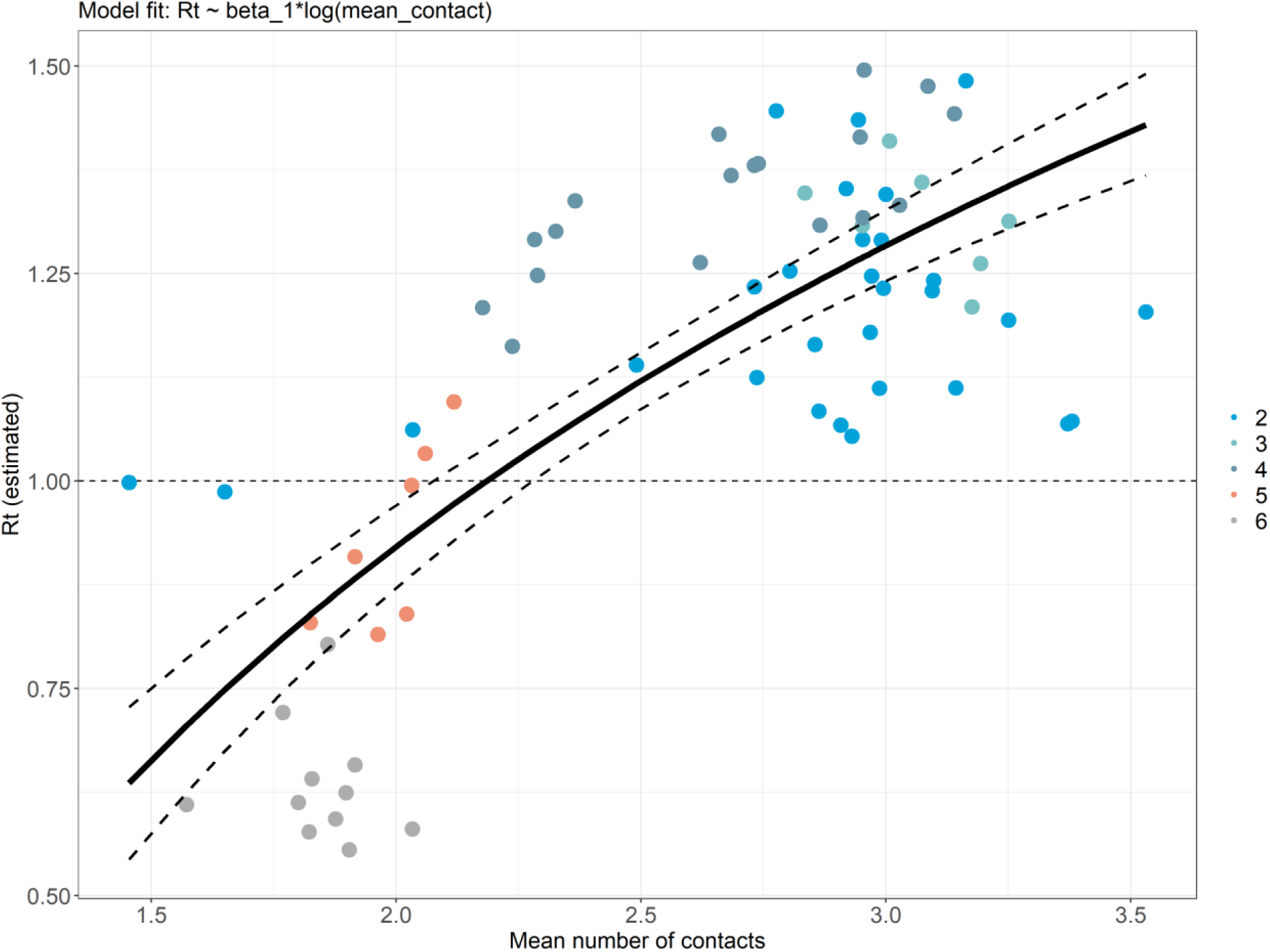
Relationship between number of contacts and reproduction number. Fitted linear regression model, regressing the instantaneous reproduction number (Rt) over the log daily mean number of contacts.

**Supp Fig 5.**
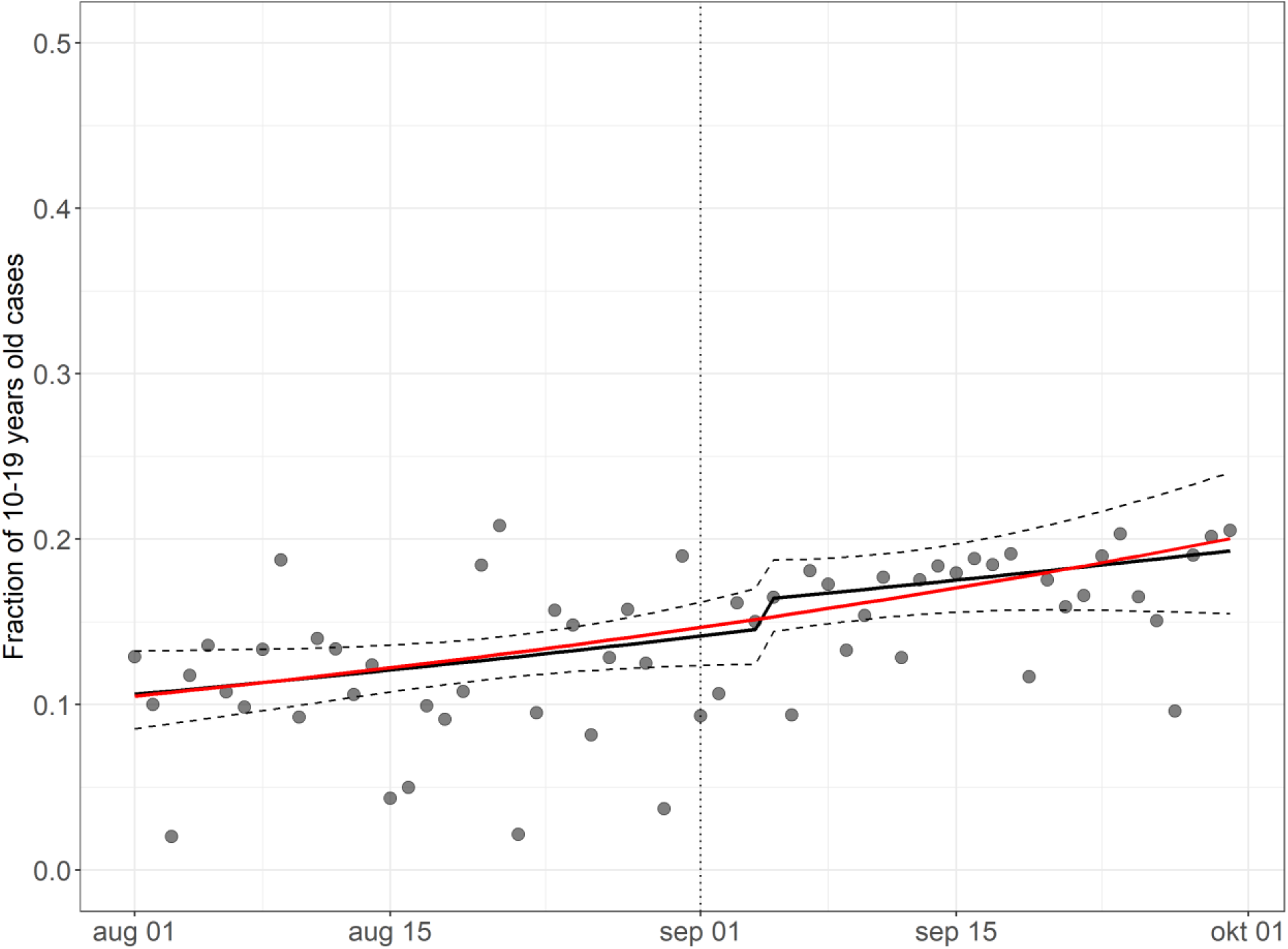
Model fit of the fraction of cases among 10-19 years old SARS-CoV-2 cases in Brussels before and after school opening, corrected for a 4-day test and report delay. Dotted line represents the timing of school opening. Red = model fit of a model assuming no step and slope change after school opening, setting variables representing weekend reporting to 0 (weekday) and number of tests among teenagers at it’s mean value. Black = model fit and 95% confidence interval of a model allowing for a step and slope change after school opening.

**Supp Fig 6:**
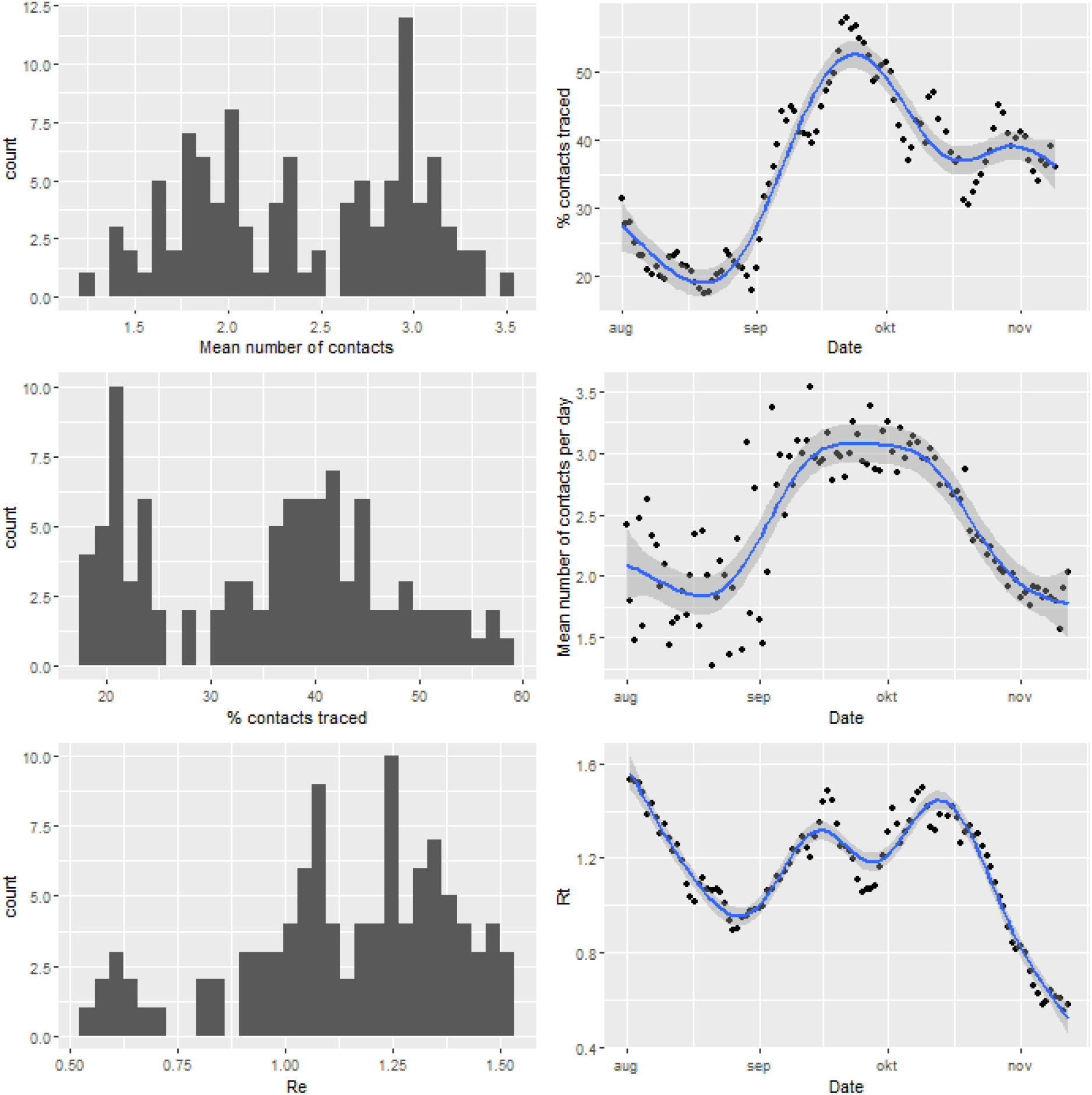
Frequency distributions of Rt, %of contacts traced and mean number of contacts.

